# Not All Entropy Is Equal: Why Representation Matters for Biological Interpretation in Dementia EEG

**DOI:** 10.64898/2026.03.09.26347355

**Authors:** Victor Edmonds

**Affiliations:** Final Stop Consulting LLC, Fuquay Varina, NC 27526, USA; Ronin Institute for Independent Scholarship 2.0, Sacramento, CA 95816, USA

**Keywords:** permutation entropy, representation, measurement validity, parameter sensitivity, dementia EEG, sensitivity analysis

## Abstract

**Objective:** To examine what a dementia-associated electroencephalography (EEG) entropy difference permits us to infer about brain dynamics by testing its dependence on permutation-entropy (PE) representation.

**Methods:** Four PE settings represented identical alpha-band data from 1,177 CAUEEG recordings. Nine duration/filtering specifications, phase reconstruction, controlled signals, age models, and 78 external participants (58 in the AD–control comparison) assessed measurement sensitivity. CAUEEG includes repeated patients without an available linkage key; its statistics describe recordings, and uncertainty estimates assume recording independence.

**Results:** In the original analysis, dementia PE contrasts ranged from *d* = −0.690 to +0.705. Across nine specifications, sub-cycle PE remained negative (*d* = −0.729 to −0.576) and coarse-sampled PE positive (+0.625 to +0.712). Cycle-spanning and high-order contrasts remained small. Phase reconstructed 90.2% of ordinal patterns; similarly high agreement occurred after spectral phase randomization. The sample-entropy contrast fell from *d* = +0.523 to +0.197 with 2 s windows on the same recordings. The external PE profile retained its ordering but did not reproduce the CAUEEG sign reversal.

**Conclusions:** The reversal is stable across the tested processing choices, but a stable contrast does not specify its biological meaning. Phase reconstruction supports a narrowband measurement account; patient-level precision and predictive generalization remain unresolved.

**Significance:** Biological interpretation requires a justified connection between the signal distinctions retained by a representation and the proposed alteration in brain dynamics.

**Highlights:**

- Changing PE representation reversed the dementia contrast on identical EEG recordings.
- The PE reversal survived processing checks; phase recovery also occurred in controlled signals.
- Repeated-patient linkage is needed for patient-level inference from CAUEEG.

## 1 Introduction

EEG entropy and complexity measures are used to investigate alterations in brain dynamics associated with dementia. EEG is inexpensive and widely available, and measures such as Lempel–Ziv complexity (LZC) and permutation entropy (PE) have shown group differences in Alzheimer’s disease (AD) and other dementias [Abásolo et al., 2006, Deng et al., 2015]; frequency-band dependence has also been demonstrated using magnetoencephalography (MEG) [Echegoyen et al., 2020]. A difference in a statistic, however, does not by itself specify which property of brain dynamics differs. Biological interpretation requires a connection between the distinctions made by the measurement and the alteration being proposed. Here we examine that connection for PE on alpha-filtered EEG.

PE represents sampled values by their rank ordering, or *ordinal pattern*, then summarizes the distribution of those patterns. Embedding order determines how many values are compared, and delay determines their spacing. At 200 Hz, order 3 with delay 1 spans 10 ms, approximately one-tenth of a 10 Hz alpha cycle; order 5 with delay 5 spans 100 ms, one complete nominal cycle. These settings preserve different portions of the waveform and define different ordinal state spaces. Methodological work has established the need to relate PE parameters to signal timescale [Riedl et al., 2013, Li et al., 2013] and finite-length sensitivity [Cuesta-Frau et al., 2019]. In dementia-related MEG, Echegoyen et al. [2020] described dominance of ascending and descending patterns at short scales and used local maxima to address it. The name “permutation entropy” therefore leaves consequential aspects of the observation unspecified: order, delay, and signal preparation remain part of what was measured.

This question arose during our own validation study. An exploratory analysis of ds004504 (*N* = 88) suggested that the alpha/theta LZC ratio was elevated in AD, prompting a larger CAUEEG analysis of prespecified LZC predictions with a corrected pipeline. PE was added as an exploratory secondary measure using the common order-3, delay-1 setting [Şeker et al., 2021]. It produced a substantial dementia contrast, approximately *d* = −0.70. Translating that setting into a 10 ms physical window prompted a further question: what signal distinction supported the apparent reduction in entropy, and what justified interpreting it as reduced biological complexity? We investigated the measurement by changing PE representation while holding the recordings, alpha-band signals, clean segments, and preprocessing fixed.

The study therefore examines how representation changes the dementia-associated contrast, what properties of the signal the resulting statistics respond to, and which interpretations remain supported after sensitivity checks. Duration and filtering variants assess whether the representation contrast persists under changes in signal preparation. Phase reconstruction, controlled signals, and spectral comparisons investigate measurement behavior; alternative measures and external data assess the scope of the result. The contribution is the quantified reversal of a clinically labeled contrast on identical recordings, its persistence across specified processing choices, and controlled-signal evidence that limits its biological interpretation. These analyses also distinguish a stable measured contrast from evidence about disease mechanisms or performance on new patients, for which additional conditions must be met.

## 2 Methods

The principal comparison changes PE representation on fixed data. Subsequent analyses address three distinct uncertainties: sensitivity to signal preparation, the signal properties recoverable from each representation, and generalization beyond the analyzed recordings. The sequence below describes the analytical rationale; prespecified objectives and exploratory follow-up are distinguished explicitly.

### 2.1 Datasets

#### 2.1.1 Discovery dataset: ds004504

The discovery dataset [Miltiadous et al., 2023] comprises resting-state EEG recordings from 88 participants: 29 healthy controls (age 67.9 *±* 5.4), 36 with Alzheimer’s disease (age 66.4 *±* 7.9), and 23 with frontotemporal dementia (age 63.7 *±* 8.2). Recordings were acquired with eyes closed using a 19-channel 10–20 montage at 500 Hz. Distributed metadata specified an A1/A2 reference. We used the original EEG files, not the provider’s ASR/ICA-processed derivatives. The dataset is publicly available on OpenNeuro.

#### 2.1.2 Principal dataset: CAUEEG

The supplied CAUEEG copy contained 1,379 annotated EEG recordings from Chung-Ang University Hospital. The source dataset comprises 1,379 recordings from 1,155 patients, including repeated visits [Kim et al., 2023]. Exclusions were 192 recordings without a qualifying diagnostic label, 7 without usable eyes-closed intervals, and 3 with less than 10 s retained after cleaning. The analysis included 1,177 recordings: 457 labeled cognitively normal (record-weighted age 65.1 *±* 9.5), 414 MCI (73.8 *±* 7.9), and 306 dementia (76.5 *±* 8.0). The retained unique-patient count is unknown: neither the distributed annotations nor the EDF patient-identification fields provided a usable linkage key. Patients with multiple visits can receive greater weight and can change diagnosis between visits. The 19 EEG channels used the 10–20 montage at 200 Hz; distributed channel labels specified an AVG reference. The dementia recordings were approximately 11.4 years older on average than normal recordings; age sensitivities were therefore examined.

The ds004504 and CAUEEG datasets were initially accessed for this research on January 9, 2026. Both were supplied in fully de-identified form, and the author had no access to information that could identify individual participants. ds004504 is openly available; CAUEEG is available through controlled academic access.

This secondary analysis was reviewed by WCG IRB and determined to be exempt under 45 CFR 46.104(d)(4). Informed consent was not applicable to the present analysis because no participants were recruited, contacted, or enrolled and only existing de-identified data were used. The ds004504 source study reports informed consent for its original collection [Miltiadous et al., 2023]. The author had no means of identifying individual participants.

### 2.2 Prespecified and exploratory analyses

Before CAUEEG was examined, the retained dataset-access request recorded an MCI-versus-control alpha/theta LZC-ratio prediction, replication of dementia-versus-control discrimination, and a comparison of MCI with dementia, with a prediction of no significant difference between those clinical groups. It also proposed examining score distributions. These documented objectives are distinguished from later changes to the processing and analysis methods; the access request was not a formal preregistration of the present pipeline. The PE and SE parameter analyses, phase and within-alpha frequency analyses, comparison with spectral power, and combined classifier were exploratory.

### 2.3 Signal processing held constant across representations

The following pipeline was applied to CAUEEG. PE parameter comparisons used the same recordings, eyes-closed intervals, artifact exclusions, band-filtered signals, and per-segment aggregation. The controlled variable was the representation defined by PE order and delay. Both cohorts retained their distributed voltage references; no common rereferencing was applied. Harmonization aligned sampling rate and estimator definitions but did not remove acquisition or reference differences.

#### 1. Eyes-closed extraction

Events were sorted, clipped to recording bounds, and compared case-insensitively after trimming labels. “Eyes Closed” began an interval; a repeated start replaced the pending start. Eyes-open, pause, recording-resumption/stop, photic, and hyperventilation-transition markers ended intervals. Each recording contributed the first eligible clean intervals up to 60 s. Mean retained data were 58.6 s (range 11.2–60.0 s), with a mean of 3.0 segments (range 1–13).

#### 2. Artifact exclusion

Artifact, movement, chewing, swallowing, speech, cough, both annotated eye-blink labels, eyelid movement, drowsiness, seizure, and sleep-attempt events were excluded with a *±*1 s buffer. Segments shorter than 2 s were discarded, including any short final fragment created by the 60 s cap. All baseline CAUEEG measure comparisons used the same retained intervals; their identities and boundaries were checked programmatically. Sensitivity variants were derived from these accepted parent intervals.

#### 3. Bandpass filtering

Alpha (8–12 Hz) and theta (4–8 Hz) signals were extracted with MNE-Python’s FIR filter (firwin design, zero-phase application) and default reflect_limited boundary padding. Each segment was filtered independently. No additional edge trim was applied; short segments and reflected boundaries remain potential sources of estimator sensitivity.

#### 4. Per-segment entropy

Entropy was computed within each segment, combined with weights proportional to segment length within each channel, then averaged equally over channels. This avoids artificial patterns at concatenation boundaries. AntroPy computed median-binarized, normalized LZC [Lempel & Ziv, 1976]. PE was the Shannon entropy of exact ordinal-pattern counts, normalized by log(*m*!) [Bandt & Pompe, 2002]. Stable ranking broke exact ties by temporal position, without adding an amplitude-dependent numerical offset; unit-scale invariance was checked explicitly. SE used a checked common implementation for all dimensions and signal lengths: distinct template pairs were counted at strict Chebyshev distance *< r*, excluding self-matches, using the same *N − m* starting positions for length-*m* and length-(*m* + 1) templates. SE was − log(*A/B*) for extension and template counts *A* and *B* [Richman & Moorman, 2000]. The principal setting was *m* = 2, *r* = 0.20 *×* SD, with population SD calculated separately within each segment and channel. No-template matches produced missing values; no-extension matches produced infinity; constant signals were explicitly flagged and assigned the continuous positive-radius limit, zero. Aggregation required every retained segment/channel estimate to be finite. The corrected full run had complete finite coverage for every reported estimate; no partially observed average was substituted.

#### 5. Spectral power

PSD was estimated independently within each unfiltered segment using Welch’s method (2 s Hann windows, 50% overlap, constant detrending). Trapezoidal integration included both endpoints of the alpha (8–12 Hz), theta (4– 8 Hz), and total (1–45 Hz) bands. Band powers were averaged over channels and weighted by segment length, without concatenating discontinuous segments. Relative alpha and theta power divided by total power; their ratio equaled absolute alpha/theta power. Identical spectral definitions were used in both cohorts.

#### 6. Spatial and ratio summaries

The primary summaries averaged entropy over all 19 channels. For the SE parameter sweep, a posterior subset (O1, O2, P3, P4, Pz, T5, T6) tested whether whole-scalp averaging diluted an alpha-localized effect. Alpha/theta ratios were computed for LZC, SE, and each PE parameterization.

The earlier ds004504 LZC analysis used 60 s clips without eyes-closed extraction or artifact exclusion. For the cross-dataset stress test described below, ds004504 was reprocessed with the harmonized pipeline.

### 2.4 PE representations in physical time

For order *m*, delay *τ*, and sampling rate *f_s_*, the embedding duration is (*m −* 1)*τ /f_s_*. Table 1 reports duration, alpha-cycle coverage, sampling geometry, and state-space size so that the observation defined by each setting is explicit. Semantic names are used in the text; software identifiers are retained for reproducibility.

**Table 1:** PE representations tested at 200 Hz. Duration is (order − 1) *×* delay *×* (1000*/f_s_*). Cycle fraction uses a 100 ms, 10 Hz reference cycle. Equal durations do not imply equal representations: order and delay determine both the sampled geometry and the number of possible ordinal states.

| Name | Code | Physical view | Order | Delay | Duration | Cycle | States |
| --- | --- | --- | --- | --- | --- | --- | --- |
| Sub-cycle | pe_o3d1 | local waveform | 3 | 1 | 10 ms | 0.1 | 6 |
| Cycle-spanning | pe_o5d5 | full-cycle span | 5 | 5 | 100 ms | 1.0 | 120 |
| Coarse-sampled | pe_o3d10 | three spaced points | 3 | 10 | 100 ms | 1.0 | 6 |
| High-order | pe_o7d3 | large state space | 7 | 3 | 90 ms | 0.9 | 5,040 |

The cycle-spanning configuration was physically motivated because its window covers one nominal alpha cycle, but the study does not establish one cycle as a uniquely correct general choice. Sub-cycle PE represents the common order-3, delay-1 setting. Coarse-sampled PE has the same total span as cycle-spanning PE but only three values separated by 50 ms. High-order PE samples a similar duration while expanding the possible state space to 5,040 patterns.

### 2.5 Phase test and comparison analyses

To examine which information the sub-cycle representation retains, we tested how closely its patterns could be recovered from the phase of the same signal. The phase account was examined analytically and on the recordings. For each clean alpha segment, instantaneous phase was obtained as *ϕ_t_* = arg*{x_t_* + *iH*[*x*]*_t_}*, where *H* is the Hilbert transform. The phase-only waveform was *u_t_* = cos *ϕ_t_*, and its ordinal patterns were computed with the same order, delay, and tie rule as the measured signal. The observed ordinal pattern for each three-sample, delay-1 vector was compared with the pattern predicted from phase alone after amplitude was discarded. Recording-level agreement and PE computed from the phase-predicted patterns were then compared with measured sub-cycle PE. Group contrasts were also computed for phase-predicted PE and for the difference between measured and phase-predicted PE.

To assess whether within-band frequency contributes information missed by relative band power, a post hoc frequency check used the 8–12 Hz spectral centroid estimated from the same unfiltered segments using 2 s Welch windows, 50% overlap, and zero-padding to a 0.1 Hz grid. Channel-averaged spectra were weighted by segment length before computing the power-weighted frequency. Zero-padding interpolates the spectrum and does not improve the underlying frequency resolution. The centroid was related to measured and phase-predicted sub-cycle PE, and age-and-group models were compared before and after adding centroid. Peak alpha frequency was a secondary check.

To compare the dependencies of alternative signal summaries, SE sensitivity was evaluated over *m ∈ {*1, 2, 3*}* and *r ∈ {*0.10, 0.15, 0.20, 0.25, 0.30*} ×* SD in whole-scalp and posterior summaries. Dimension *m* = 1 compares scalar-template matches with their two-sample extensions and is a valid low-dimensional comparison. The implementation was checked against independent brute-force pair counts and across the 5,000-sample library branch boundary that had caused missing estimates in the earlier implementation. LZC, SE, and relative spectral power supplied comparison representations.

To assess whether the measured contrasts transferred to another cohort, the original ds004504 recordings were downsampled from 500 to 200 Hz with MNE-Python anti-aliased resampling. One-second epochs were flagged when any retained channel exceeded 150 *µ*V peak-to-peak or five times the median channel–epoch variance. Channels flagged in more than half the epochs were removed; at least ten channels were required. Consecutive good epochs were merged; spans shorter than 2 s were discarded. At least 20 s of usable data were required, with the same 60 s cap and segment-level estimators as CAUEEG. This retained 78 participants (31 AD, 27 control, 20 FTD); the AD–control contrast used 58. The source acquisition was eyes closed, but equivalent CAUEEG-style event-marked segmentation was unavailable. Identical integer PE parameters now represented identical physical durations, while acquisition/reference differences and possible severity or composition differences remained. Because ds004504 had also informed the initial discovery work, this was an external portability stress test rather than untouched validation. Retained-sample characteristics are reported in Supplementary Section S8.

### 2.6 Statistical analysis

The primary estimands are descriptive, record-weighted contrasts within CAUEEG and participant-level contrasts externally. Cohen’s *d* is disease minus reference, using the sample-size-weighted pooled sample SD. Descriptive single-measure AUC was direction-corrected as max(AUC, 1 − AUC) after observing direction; model-test AUC was not direction-corrected. The retained CAUEEG Welch tests, bootstrap intervals, and HC3 regressions assume independent recordings. Because repeated patients cannot be grouped, these quantities are conditional summaries and do not establish patient-level precision. Age or multiple-testing adjustment does not repair that dependence. Age checks used outcome residualization across all CAUEEG recordings and restriction to ages 70–80 (*n* = 508 recordings: Normal 129, MCI 230, Dementia 149); restriction is not individual matching.

For transparency, earlier conditional inferential tables are retained. Benjamini–Hochberg correction covered 42 distinct default/sweep measures per CAUEEG diagnostic contrast, 13 defaults externally, 26 measure-by-age-approach tests per CAUEEG contrast, 13 quadratic-age tests, four phase contrasts, 12 frequency correlations, and three documented LZC-objective contrasts. Tables distinguish raw *p* from adjusted *q*; nominal frequency/interaction coefficients are exploratory. Original pointwise 95% intervals used 1,000 recording resamples within CAUEEG groups and participant resamples externally. The primary PE table emphasizes descriptive effect sizes. Original logistic models used training-fold standardization and L2 regularization in stratified recording-level 10-fold cross-validation repeated ten times. Repeat-mean percentile ranges describe split variability only. Repeated patients may cross folds, and full-cohort feature selection preceded validation; these results cannot establish performance on new patients. Interaction coefficients came from separate unpenalized models with the same dependence limitation.

### 2.7 Signal and model sensitivity analyses

To separate representation sensitivity from duration and filtering effects, an exploratory protocol fixed nine specifications before inspection of the complete sensitivity tables: the original segments; 2 s windows filtered individually or selected after filtering their clean parent segment; parent-filtered 4, 8, and 16 s windows; paired 2 s interior-window variants; and exactly ten parent-filtered 2 s windows. Windows never crossed clean-segment boundaries. Interior windows lay at least 165 samples (0.825 s) inside each parent boundary. Eligibility, retained duration, and original estimates on the same eligible recordings were tracked. Ordinal-count pooling within channel/recording was compared with mean segment entropy without constructing cross-window patterns; it changes the estimand. Sensitivity summaries required at least 20 eligible records per diagnostic group. Conditional paired bootstrap summaries used 2,000 draws; the 17-measure BH families did not correct across specifications. These checks were not preregistered (Supplementary Section S5).

Controlled frequency/length simulations and Fourier-phase randomization examined whether the observed measurement behavior could arise under simpler signal constructions. A seeded probe used 20 CAUEEG recordings per group and all 58 external AD/control participants, with ten surrogates per recording, preserving each parent segment/channel’s Fourier magnitudes. These were descriptive probes, not formal nonlinearity tests. Additional age models, including group-specific splines, assessed sensitivity to the form of the age relationship. To examine predictive transfer separately from descriptive separation, the provider’s no-overlap training and test partitions supplied an exploratory check, using training-only scaling and fixed logistic settings. Those files remove training-patient overlap from test, but do not identify unique patients within each partition; feature development had already used pooled data (Supplementary Sections S4 and S7).

## 3 Results

### 3.1 Representation changes the dementia contrast

Changing only the PE representation changed the dementia effect. Applying the four settings to the same 1,177 recordings, alpha-band signals, and clean segments yielded dementia-versus-normal effect sizes ranging from *d* = −0.690 to *d* = +0.705 (Table 2; Figure 1). Sub-cycle PE was lower in dementia, coarse-sampled PE was higher, and the cycle-spanning and high-order configurations had small observed contrasts. MCI comparisons had the same ordering at smaller magnitude.

**Figure 1:**
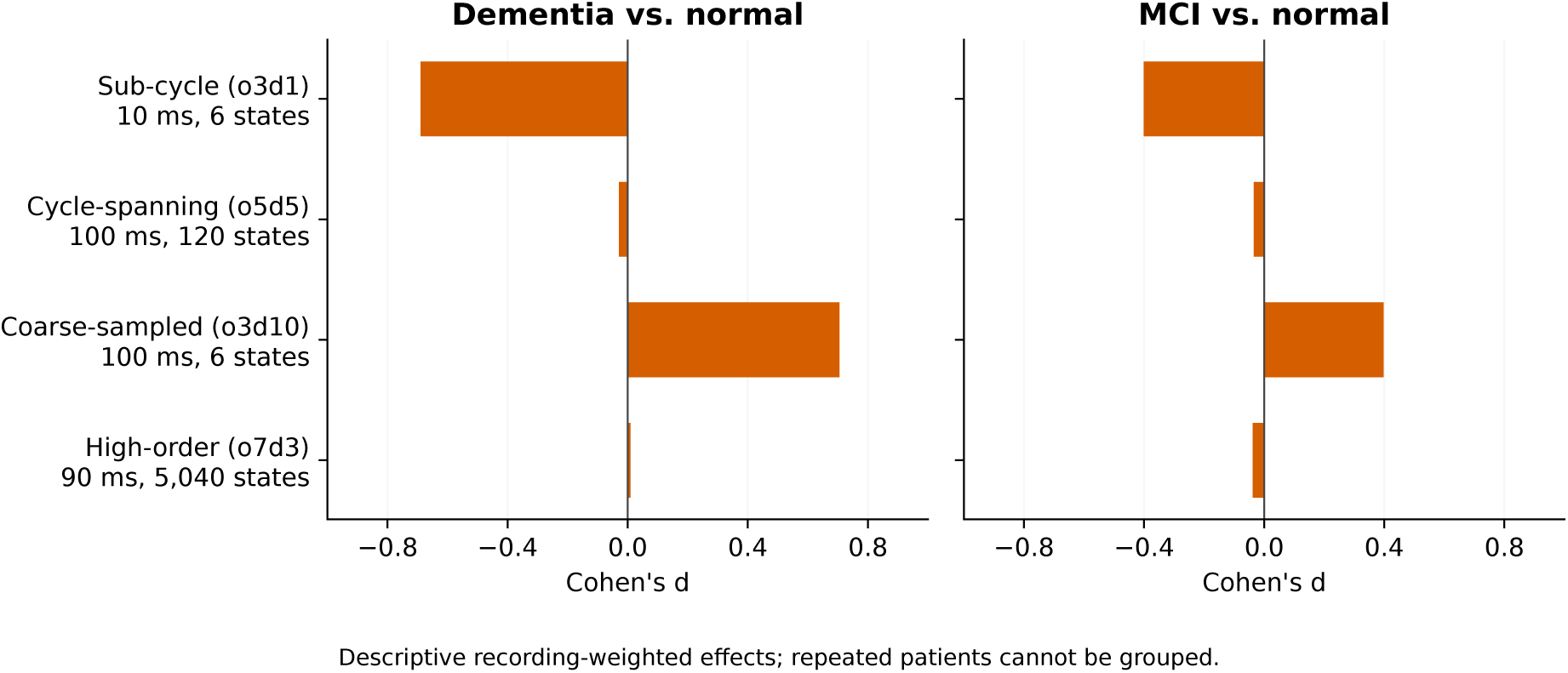
Changing PE representation reverses the measured dementia contrast. Cohen’s *d* for dementia versus normal (left) and MCI versus normal (right) across four configurations applied to identical alpha-band data. Panels share the same horizontal scale and show descriptive point estimates. Conditional recording-bootstrap intervals are retained in the aggregate supplementary tables; they do not establish patient-level precision. The negative, positive, and near-null effects establish parameter sensitivity; they do not by themselves explain its mechanism.

**Table 2:** Descriptive alpha-band PE contrasts in CAUEEG (*N* = 1,177 recordings). Identical recordings and preprocessing; only the representation changes. AUC is direction-corrected after observing direction. Repeated patients are not grouped; these are record-weighted effects. Conditional tests are retained in Supplementary Section S1.

| Representation | $d_{\text{Dem}}$ | $d_{\text{MCI}}$ | $\text{AUC}_{\text{Dem}}$ | $\text{AUC}_{\text{MCI}}$ |
| --- | --- | --- | --- | --- |
| Sub-cycle PE | −0.690 | −0.401 | 0.699 | 0.613 |
| Cycle-spanning PE | −0.029 | −0.034 | 0.514 | 0.512 |
| Coarse-sampled PE | +0.705 | +0.398 | 0.701 | 0.611 |
| High-order PE | +0.009 | −0.038 | 0.504 | 0.517 |

The common measurement name encompassed different observations of the waveform. The two order-3 settings reversed direction while retaining the same six possible patterns; the two 100 ms settings produced different contrasts despite equal duration. We next tested whether the opposing directions persisted under changes in signal preparation.

### 3.2 Opposing directions persist across processing choices

Across all nine specifications, dementia sub-cycle PE remained negative (*d* = −0.729 to −0.576), and coarse PE positive (+0.625 to +0.712). Cycle-spanning and high-order group contrasts remained small (Supplementary Section S5). Matching the eligible recordings to their original estimates separated processing changes from changed eligibility. Excluding the few records with constant-window flags preserved the direction reversal. All original default/phase estimates reproduced, and no included-record estimate was nonfinite.

Joint age models retained opposite dementia PE directions, including group-specific spline curves standardized over the observed 70–80 age distribution (sub-cycle −0.611 and coarse +0.320, in full-cohort outcome SD). MCI was less stable: the corresponding coarse contrast was +0.046 and its effect changed sign in the small official test partition. These record-level models do not eliminate patient dependence or unmeasured confounding (Supplementary Sections S2 and S7).

These checks establish persistence of the dementia representation contrast under the tested choices. They leave open which waveform properties support each statistic and how those properties relate to disease.

### 3.3 Sub-cycle patterns are closely recoverable from phase

The phase analysis asked how much of the sub-cycle observation remained recoverable after amplitude was discarded. Figure 2 shows the physical difference: the sub-cycle configuration sees three adjacent samples on a small arc, whereas the cycle-spanning configuration samples across a complete nominal oscillation.

**Figure 2:**
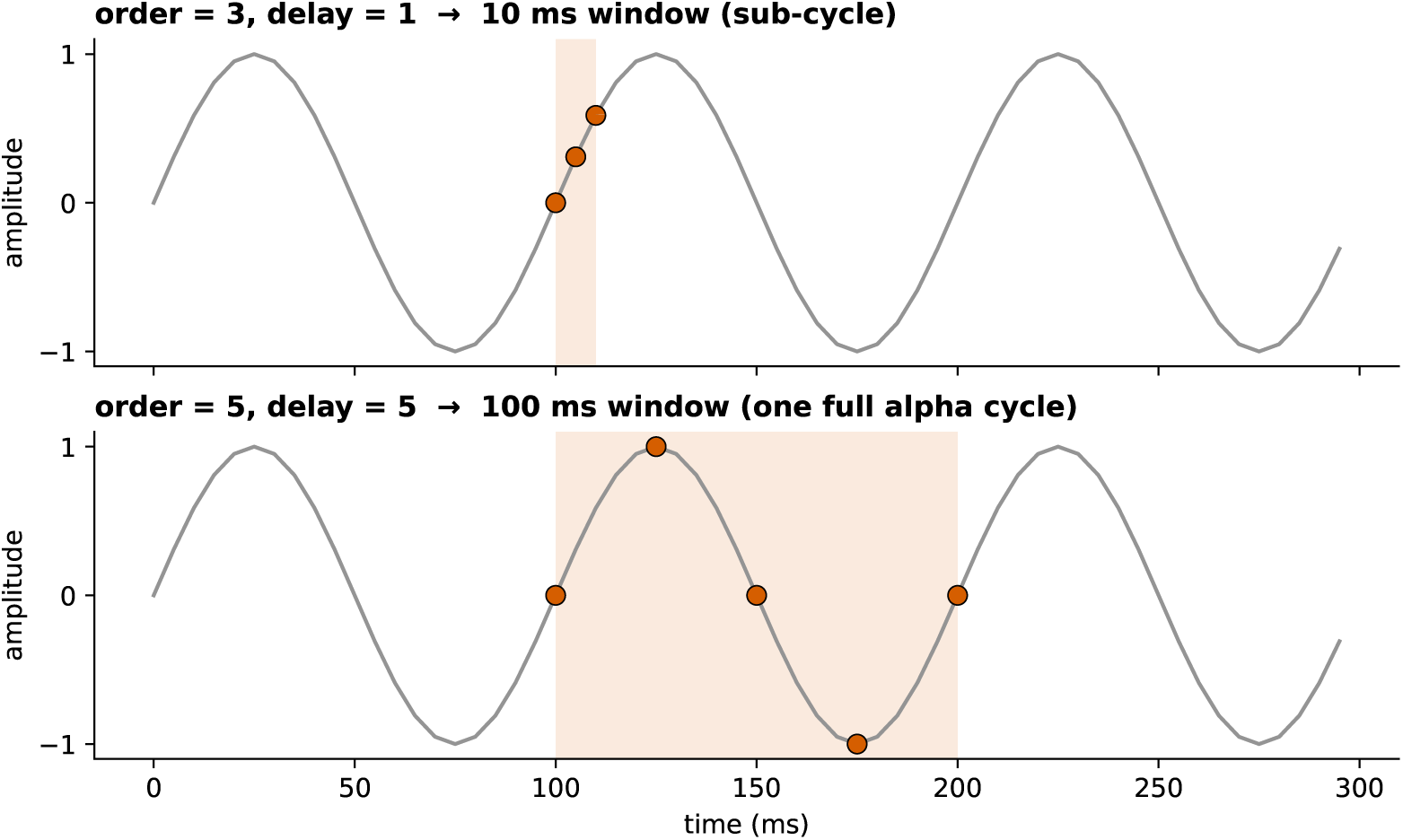
The two representations see different portions of an alpha waveform. A synthetic 10 Hz signal sampled at 200 Hz is shown with the 10 ms sub-cycle window (top) and 100 ms cycle-spanning window (bottom). The schematic explains the physical scale of the comparison; it does not establish one window as universally optimal.

For a narrowband signal *x*(*t*) *≈ A*(*t*) sin(*ωt* + *φ*_0_), three consecutive values can be written

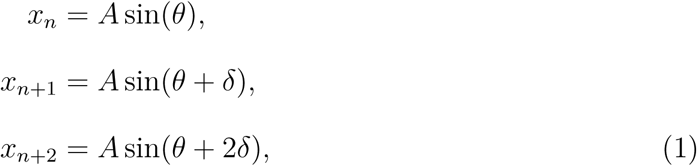

where *θ* is phase in the sine convention (*θ* = *ϕ* + *π/*2 relative to the Hilbert-phase cosine convention) and *δ* = 2*πf/f_s_*. At *f* = 10 Hz and *f_s_* = 200 Hz, *δ* = *π/*10. When amplitude changes little across the three samples, their rank ordering depends on *θ*. The six possible order-3 patterns partition the phase circle into sectors.

Those sectors are unequal. For uniformly distributed phase at 10 Hz, the rising and falling patterns each occupy 45% of the circle, while four turning-point patterns occupy 2.5% each. The resulting normalized PE is approximately 0.61 rather than 1.0. This is a uniform starting-phase ensemble calculation. A single noiseless 10 Hz orbit sampled at 200 Hz revisits only 20 phases and need not have that ensemble entropy. Modulation, within-band frequency variation, and phase resets can alter the estimate.

The recording-level reconstruction closely approximated the sub-cycle patterns and entropy. Phase alone predicted a mean 90.2% of observed ordinal triplets across 1,177 recordings. Phase-reconstructed PE closely tracked measured sub-cycle PE (Pearson *r* = 0.983, Spearman *ρ* = 0.998; Figure 3). High agreement is expected because Hilbert phase is derived from the same narrowband signal; the empirical purpose was to quantify the degree of recovery from phase. Phase-reconstructed PE retained a similar dementia contrast (*d* = −0.701, versus −0.690 for measured PE); the measured-minus-reconstructed residual contrast was small and opposite (*d* = +0.180).

**Figure 3:**
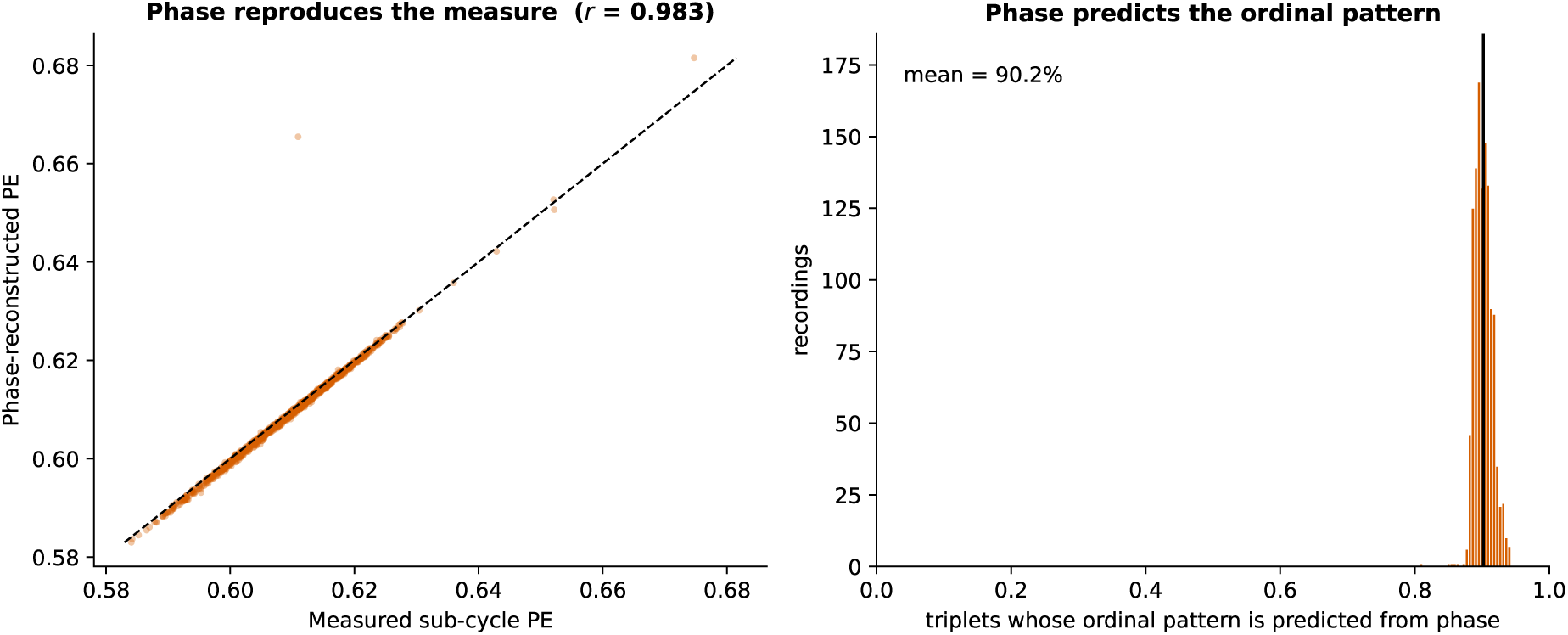
Phase geometry reproduces sub-cycle PE in the recordings. Left: measured sub-cycle PE against PE recomputed from Hilbert phase alone, with amplitude discarded (*r* = 0.983; dashed line is identity). Right: recording-level proportion of triplets whose ordinal pattern is predicted from phase (mean 90.2%). High agreement is expected for Hilbert phase derived from the same narrowband signal; the result quantifies recovery from phase for this representation. It does not imply that broadband PE or PE in general reduces to phase.

The phase result also clarifies why weak correlation with relative alpha power (*r* = −0.136) does not establish independence from spectral structure. In a post hoc check, the 8–12 Hz centroid correlated strongly with measured sub-cycle PE (Pearson *r* = 0.853, Spearman *ρ* = 0.906) and phase-reconstructed PE (Pearson *r* = 0.831, Spearman *ρ* = 0.907). Adding centroid to an age-and-group model attenuated the standardized dementia coefficient by 56.3%, from −0.454 to −0.198. The association attenuated after including within-band frequency; the change is model-dependent and is not a causal mediation fraction. Restricting centroid to 8–12 Hz does not capture slowing below the fixed band; these associations do not establish causation.

In the CAUEEG probe, pattern agreement was 90.24% in the recordings and 90.13% after spectral phase randomization. Controlled alpha-filtered noise also showed high agreement. These comparisons show that close reconstruction is compatible with a general narrowband property; they do not establish disease-specific nonlinear dynamics or a unique explanation of the group contrast (Supplementary Section S6).

### 3.4 Sampling and estimation affect other PE representations

Coarse-sampled PE produced a positive dementia effect (*d* = +0.705), opposite to the sub-cycle effect. The two order-3 measures were weakly correlated (*r* = −0.126) despite sharing the same six-state space.

One plausible contributor is frequency-sensitive sampling. A delay of 10 at 200 Hz compares values at 50 ms intervals, corresponding to a 20 Hz comparison rate with a 10 Hz Nyquist boundary inside alpha [Popov et al., 2013]. In controlled noisy sinusoidal sweeps, coarse PE had a trough around 10 Hz while sub-cycle PE rose with frequency. Slowing below 10 Hz can therefore produce opposite changes in the two settings. This demonstrates plausibility under known generators, not a uniquely identified explanation for the clinical contrast (Supplementary Section S6).

High-order PE, the 90 ms configuration with 5,040 possible states, had a near-zero original contrast (*d* = +0.009) and closely correlated with cycle-spanning PE (*r* = 0.986). Guidance calls for *N ≫ m*! for PE estimation [Cuesta-Frau et al., 2019]. Here estimation was performed separately on segments as short as 400 samples, before averaging; the 60 s recording cap therefore overstates the length available to many individual estimates. The small group contrast persisted under the tested duration and pooling choices, although absolute high-order entropy was length-sensitive. It does not establish equivalence or a mechanistic absence of a dementia association.

### 3.5 Alternative measures have different dependencies

SE, LZC, and spectral power supplied comparisons for evaluating what an entropy contrast adds and what other dependencies it introduces. Each summarizes different distinctions in the same signal.

SE was sensitive to dimension and tolerance. At *r* = 0.20 *×* SD, whole-scalp alpha-band effects were *d* = +0.042 for *m* = 1, *d* = +0.523 for *m* = 2 (AUC 0.722), and *d* = +0.871 for *m* = 3. The whole-scalp *m* = 2 grid ranged from *d* = 0.450 to 0.575; its minimum inter-tolerance correlation was 0.739. Posterior restriction gave *d* = +0.620 at *m* = 2*, r* = 0.20. Complete finite coverage was confirmed across the grid. Short-segment finite-sample effects remain possible, and complete coverage does not establish absence of estimator bias. Supplementary Section S3 reports the full sensitivity results.

The descriptive LZC-ratio directions partly aligned with the documented objectives: the ratio was elevated relative to normal in dementia (*d* = +0.473, AUC 0.624) and MCI (*d* = +0.229), and the dementia direction was also present in ds004504 AD. MCI and dementia differed (*d*_MCI−Dem_ = −0.270), whereas the documented objective predicted a non-significant comparison. The recording-level tests cannot settle that patient-level prediction. A non-significant difference would not itself have established equivalence.

The spectral baseline remained informative. Relative alpha/theta power was lower in dementia (*d* = −0.760, AUC 0.754), and relative theta power had *d* = +0.783 (AUC 0.709). AUC ranking is restricted to the default measures in Supplementary Section S1: the exploratory SE grid reached apparent AUC 0.799, so power was not the best over all explored parameter settings. The LZC ratio shared 47.6% of its variance with the power ratio (*r* = −0.690). SE*_α_* was weakly correlated with relative alpha power (*r* = −0.056); this describes little linear association and does not establish spectral independence or incremental disease-relevant information. Figure 4 summarizes the default comparison.

**Figure 4:**
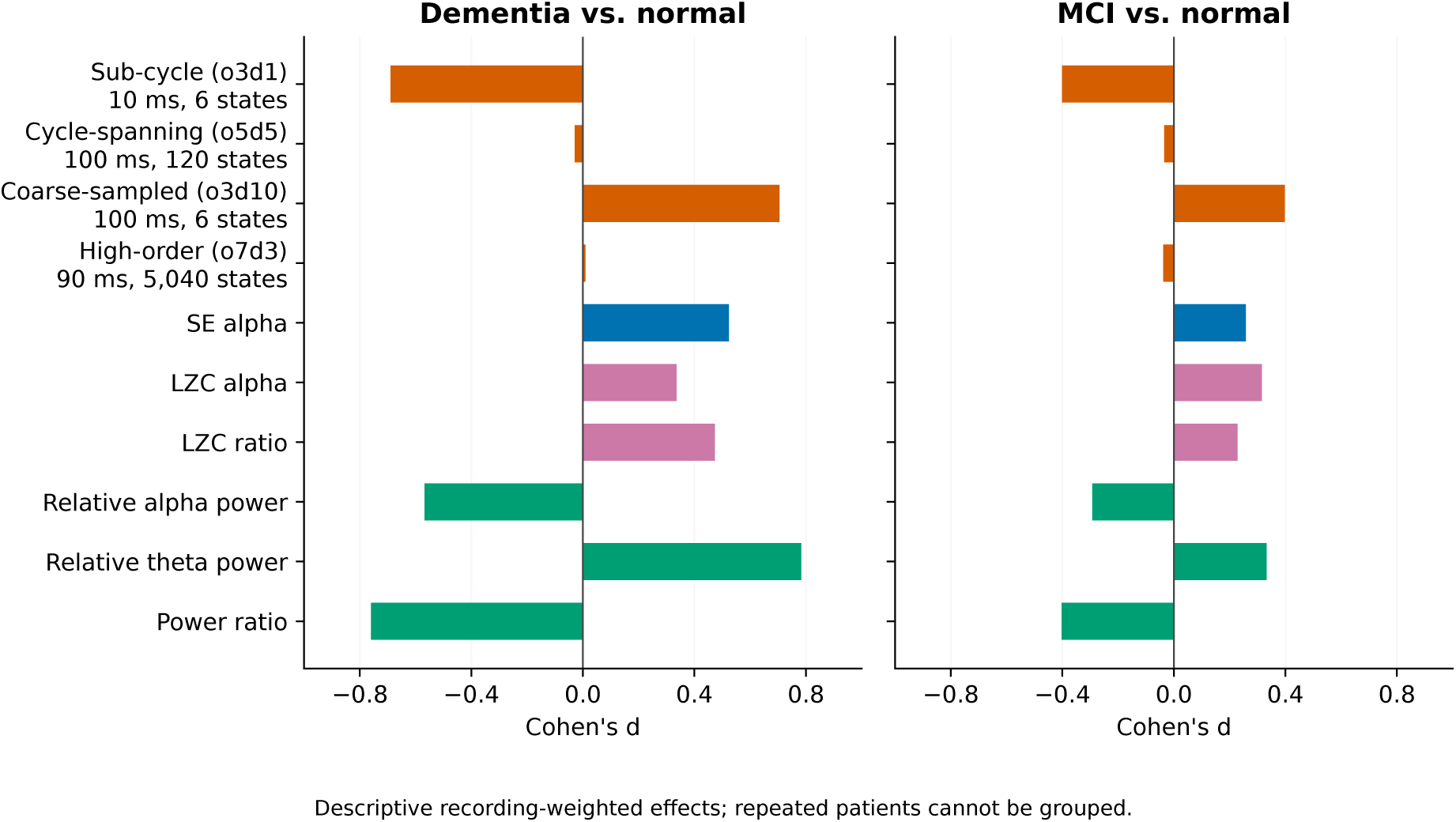
Entropy, complexity, and spectral representations retain different group contrasts. Bars show descriptive Cohen’s *d* for dementia versus normal (left) and MCI versus normal (right), on the same horizontal scale. Relative theta power had the largest raw dementia effect; relative alpha/theta power is shown as a spectral benchmark. Repeated patients are not grouped; conditional intervals are retained in the aggregate supplementary tables and do not establish patient-level precision. The exploratory SE parameter grid is reported separately. This within-cohort comparison is a spectral benchmark, not evidence that power is a sufficient dementia biomarker.

**Figure 5:**
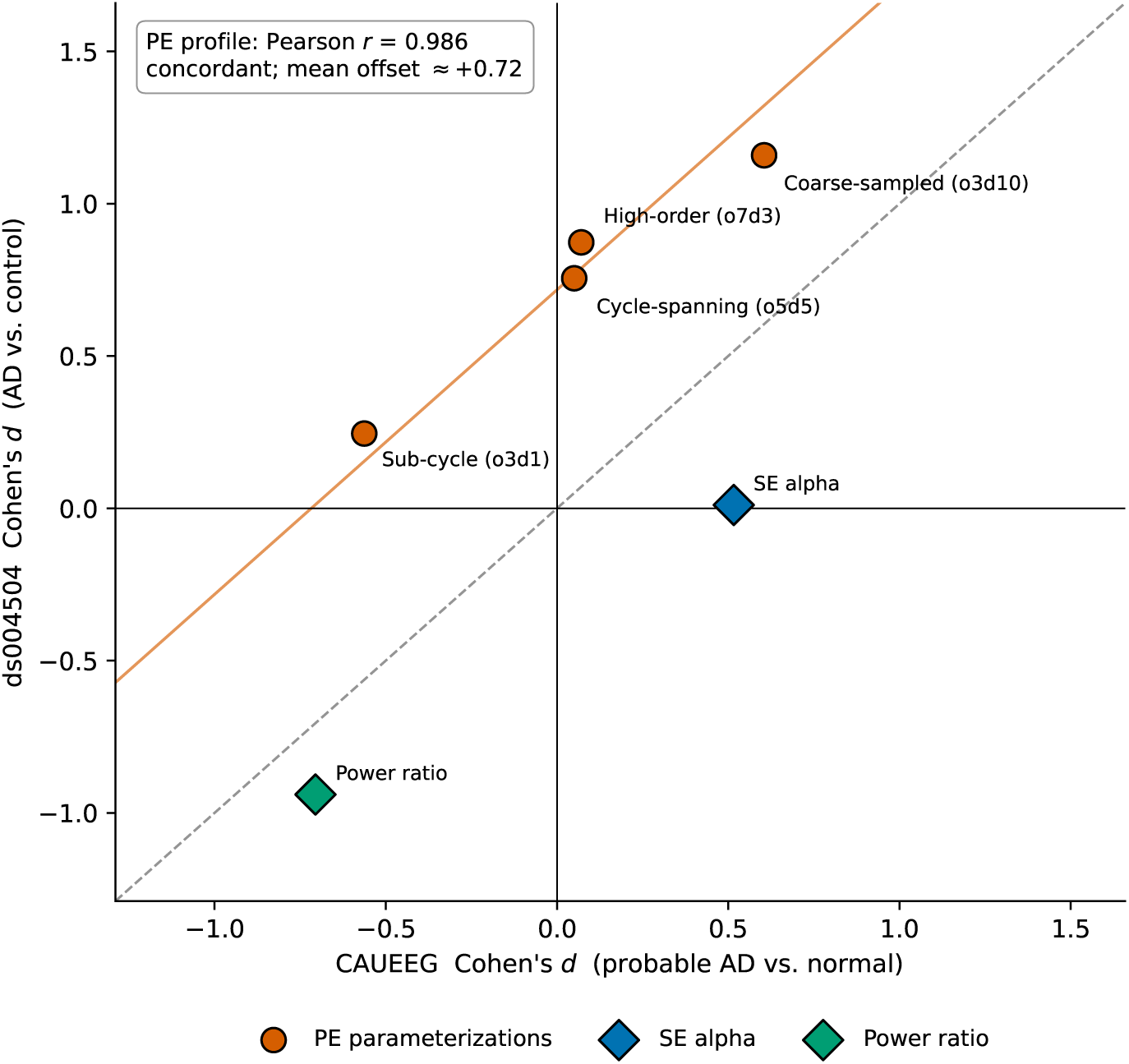
Parameter response can be concordant while biomarker output is not portable. Cohen’s *d* in CAUEEG probable AD versus normal is plotted against harmonized ds004504 AD versus control. The four PE configurations (circles) follow a similar ordering but are shifted upward; the reported *r* = 0.986 is based on four points. The dashed line is identity, and the orange line shows the mean offset in standardized PE effects; this is not an offset in absolute entropy units. Under the original segment specification, SE (diamond) has a near-zero external estimate, whereas relative alpha/theta power retains its negative direction. Differences between cohorts prevent attribution of the offset to one cause.

SE magnitude was less stable: the original *d* = +0.523 fell to +0.197 with parent-filtered 2 s windows on the same 306 dementia and 457 normal recordings, and to +0.120 with interior windows. Removing constant-window records reduced SE further in some specifications. The LZC-ratio direction remained positive but its magnitude varied; power-ratio effects remained negative. These comparisons support representation sensitivity while limiting claims based on SE’s original magnitude.

### 3.6 External comparisons and exploratory prediction

The two cohorts did not support a portable PE group effect. The four configurations had a concordant response pattern (Pearson *r* = 0.986, Spearman *ρ* = 1.000), displaced by a mean upward offset of 0.717 in pooled-SD Cohen’s *d*. This four-point correlation is descriptive and does not establish a general invariant.

Sub-cycle PE changed from *d* = −0.563 in CAUEEG probable AD versus normal to *d* = +0.245 in ds004504 AD versus control. Cycle-spanning PE changed from *d* = +0.050 to *d* = +0.755 (external *p* = 0.008, *q* = 0.018). SE was positive in CAUEEG (*d* = +0.516) but had a near-zero original estimate in ds004504 (*d* = +0.011, *p* = 0.966, *q* = 0.966). These results do not support an unqualified portable entropy biomarker under the tested processing.

Relative alpha/theta power retained a negative direction (CAUEEG probable AD *d* = −0.705; ds004504 *d* = −0.940). The LZC ratio retained a positive direction (*d* = +0.523 and +0.495, respectively), although its shared variance with power limits an independent complexity interpretation. The probable-AD-only CAUEEG check (*n* = 226 recordings) preserved the within-cohort PE parameter pattern. Severity, recording state, sample size, and composition remain possible contributors to the offset.

In the provider’s no-overlap test partition (22 dementia, 35 normal recordings), the fixed model gave AUC 0.740, versus 0.742 for SE alone and 0.690 for power. This small exploratory check did not establish an advantage for the combination. The original recording-level cross-validation, full results, and partition limitations are in Supplementary Section S4.

These comparisons distinguish separation within the analyzed recordings from performance in other patients. The unavailable repeated-patient linkage limits uncertainty and validation even where a descriptive direction persists.

## 4 Discussion

The initial dementia contrast motivated an investigation of what PE had observed. On identical alpha-band data, changing representation reversed that contrast, and the opposing directions persisted across the tested signal specifications. Phase and controlled-signal analyses then clarified properties to which the measurement responds. The resulting contribution is a description of how representation changes a clinically labeled EEG contrast, with evidence delimiting its interpretation. Processing stability, biological meaning, and patient-level generalization require separate justification.

### 4.1 Representation is part of the biological inference

The same alpha-band EEG supported lower, higher, and near-null PE effects when only order and delay changed. The parameters therefore did more than tune effect magnitude: they changed the physical question. A 10 ms window samples local alpha-waveform geometry; a 100 ms window spans a nominal cycle; three widely spaced values over 100 ms define yet another representation. Duration makes the temporal scale explicit, while order and delay also determine sampling geometry and the ordinal state space. A biological interpretation must account for those distinctions.

Different representations can legitimately produce different contrasts because they preserve different properties. Agreement across settings is therefore not a universal criterion of validity, and the present comparison does not select one optimal PE parameterization. The requirement is to connect the chosen observation to the biological question. A finding described simply as “lower entropy in dementia” omits the representation on which its direction depends; that omission matters if the reader carries it forward as a general statement about reduced brain complexity.

### 4.2 Signal reconstruction clarifies measurement behavior

Recovery from phase identifies information sufficient to closely approximate the sub-cycle statistic on these signals. High agreement after spectral phase randomization and in controlled filtered noise shows that this behavior can arise without a disease-specific generator. Together with the within-alpha centroid association, these observations support a general narrowband measurement account. They do not identify a unique cause of the clinical group contrast or a causal fraction attributable to spectral slowing.

The controlled sweeps support frequency-sensitive sampling as a plausible contributor to the coarse reversal. For high-order PE, length curves and count pooling show that absolute entropy can change while the observed group contrast remains small. The simulations are not disease models, and neither account uniquely identifies the clinical mechanism.

### 4.3 Added information and generalization

PE, SE, LZC, and power discard different information. Ordinal PE removes distances between values but can still respond when irregularity changes rank patterns. SE evaluates recurrence within a tolerance-scaled distance; its effects depended on embedding dimension and tolerance, and its positive CAUEEG contrast was not reproduced under the tested external specifications. This does not establish an absent effect or a significant difference between cohorts. SE was weakly linearly associated with relative alpha power in this sample. This does not establish spectral independence or incremental disease-relevant information. Broadband EEG sample entropy has likewise been shown to track the log ratio of fast to slow spectral power across sleep and aging [Bruce et al., 2009], reinforcing the need to test more than one spectral summary.

LZC retained the same descriptive direction across cohorts, but 47.6% of the variance in its alpha/theta ratio was shared with the spectral power ratio. The power ratio provided a strong default-measure benchmark and retained direction across cohorts, consistent with established EEG spectral slowing in dementia [Jeong, 2004]; this does not make it a complete disease model. It makes spectral slowing a necessary baseline. A more elaborate measure should be evaluated for what it adds beyond that baseline and whether the added information transfers.

The available comparisons did not validate a portable cross-cohort entropy biomarker. PE’s four-setting response pattern was concordant but shifted in standardized effect size; SE had a near-zero original external estimate that shifted under duration controls; LZC retained direction but was substantially spectral. These outcomes caution against using within-cohort statistical strength as a substitute for measurement interpretation or external stability.

### 4.4 What a subsequent study needs to establish

For a band-filtered entropy contrast to support a biological interpretation, the observation procedure must remain visible in the claim. Report the filtering, embedding order and delay, physical duration, state space, and aggregation rule. Test alternatives that address the proposed interpretation and describe which conclusions depend on those choices. Controlled signals and spectral comparisons can then assess whether simpler waveform properties reproduce the relevant behavior. The goal is to establish what the chosen representation contributes to the neurological question.

Generalization requires evidence beyond that measurement account. Patient linkage is needed to define visit weighting, estimate patient-level uncertainty, and separate patients during validation. External comparisons should examine whether the interpreted signal property and the diagnostic contrast transfer under specified recording conditions. A concise report should preserve what was held fixed, what changed, what was observed, and the scope of the inference. In this study, a substantial initial contrast supplied the reason to investigate; its magnitude alone did not settle what it meant.

### 4.5 Limitations

CAUEEG’s repeated patients are a material limitation. The retained unique-patient count is unknown, and unequal visit counts can affect point estimates as well as precision. Welch tests, bootstrap intervals, and HC3 models treating recordings independently do not account for within-patient dependence, including visits with different diagnoses. The available no-overlap benchmark partitions do not deduplicate the pooled data. Patient linkage is needed for a prespecified visit-selection or patient-cluster analysis and patient-grouped validation. The present central claim is therefore about representation-dependent contrasts in these recordings.

The LZC objectives were documented before CAUEEG analysis, whereas PE, SE, phase, spectral comparisons, and classifier analyses were exploratory. The external cohort was small (58 AD and control participants after harmonization) and had already informed discovery. Comparable disease-severity measurements were unavailable across cohorts. Although both analyses targeted eyes-closed EEG, equivalent event-marked segmentation was unavailable externally and voltage references were not made identical. Possible severity differences, acquisition/reference conditions, sample size, and composition therefore cannot be separated, and the four-point PE profile correlation should not carry inferential weight beyond the tested configurations.

The analysis is restricted to fixed 8–12 Hz alpha and 4–8 Hz theta bands. Individual slowing may move a dominant rhythm outside those boundaries. Results may differ for broadband PE, other bands, sampling rates, weighted PE, or dispersion entropy [Deng et al., 2015, Azami et al., 2026]. Controlled generators illustrate sampling/estimation behavior but cannot identify the full mechanism in clinical EEG. Long and interior windows left too few external participants for several planned contrasts; those specifications provide no whole-cohort external robustness claim.

CAUEEG contains mixed dementia etiologies and a clinical-reference normal group rather than community controls. Medication data were unavailable. Cholinesterase inhibitors, benzodiazepines, antiepileptic drugs, and other psychotropic medications can alter EEG spectra and entropy, but their net contribution cannot be resolved here. Retrospective clinical recordings also vary more than research-grade acquisitions.

Sex and gender were not included in the fitted models. The source-labeled Gender field was available for ds004504 and is summarized descriptively for retained participants in Supplementary Section S8; a comparable field was unavailable in the supplied CAUEEG annotations. Effect modification and adjustment for sex or gender were not evaluated.

Age differed substantially between diagnostic groups. Flexible age models and age restriction assess sensitivity but do not establish adequate causal adjustment. MCI effects were less stable than the principal dementia comparison. The combined classifier lacks independent validation and did not establish an advantage over SE alone in the exploratory provider-split check. Clinical thresholds and incremental clinical utility are not established.

## 5 Conclusion

On these alpha-band recordings, changing PE representation reversed the contrast between dementia and normal recordings. That reversal persisted across the tested duration and filtering choices. Phase reconstruction and controlled signals support a measurement-geometric interpretation of sub-cycle PE. SE effect magnitude was sensitive to window duration; LZC retained direction while sharing substantial spectral variance; and relative alpha/theta power provided a consistent directional baseline. Patient-level precision and predictive generalization remain unresolved without repeated-patient linkage.

A biological interpretation requires a justified connection between the waveform distinctions retained by the chosen representation and the proposed alteration in brain dynamics. Making that connection explicit preserves what the measurement establishes and identifies the evidence needed for the next inference.

## Data Availability

All data produced are available online.
Analysis within this manuscript:
https://doi.org/10.5281/zenodo.18789520
ds004504 dataset:
https://openneuro.org/datasets/ds004504
CAUEEG dataset (requires application)
https://doi.org/10.1016/j.neuroimage.2023.120054

https://doi.org/10.5281/zenodo.18789520

https://openneuro.org/datasets/ds004504

https://doi.org/10.1016/j.neuroimage.2023.120054

## Data and Code Availability

The ds004504 dataset is available on OpenNeuro (https://openneuro.org/datasets/ds004504; v1.0.8, DOI: 10.18112/openneuro.ds004504.v1.0.8). The CAUEEG dataset is available for academic use under controlled access, granted on application to the data providers rather than by open download; access procedures are described by Kim et al. [2023]. Analysis code and the pinned environment are publicly available at https: //github.com/Final-Stop-Consulting/caueeg-entropy; the code version for this revision is commit 81a02cb, archived as release v3.0.1 at Zenodo (DOI: 10.5281/zenodo. 23021214). The accompanying Supplementary_Results.zip contains aggregate statistical tables, controlled-signal results, a data dictionary, and a provenance manifest for the reported analyses. Source EEG and record-level clinical data remain with the respective data providers.

## Contributors

Victor Edmonds: Conceptualization, methodology, software, formal analysis, investigation, data curation, visualization, writing–original draft, writing–review and editing, and project administration. The author accepts responsibility for the final content.

## Declaration of generative AI use

During the preparation of this work the author used Claude (Anthropic) and Codex (OpenAI) to assist with code implementation, computational auditing, and LaTeX formatting. The author is responsible for reviewing and verifying the analyses and final manuscript.

## Ethics Statement

This study was reviewed by WCG IRB and determined to be exempt under 45 CFR 46.104(d)(4) as secondary research using existing de-identified data. Informed consent was not applicable to the present secondary analysis. The ds004504 source study reports informed consent for its original data collection [Miltiadous et al., 2023]. No participants were recruited, contacted, or enrolled, and the author had no means of identifying individual participants. IRB Tracking Number: 20260808; Protocol Number: FSC-2026-001; Board Action Date: March 5, 2026.

