## Supplemental Data for "Not All Entropy Is Equal: Why Representation Matters for Biological Interpretation in Dementia EEG"

Victor Edmonds

**Purpose of the supplementary analyses.** These analyses distinguish a persistent measurement result from evidence about mechanism or new-patient performance. Sections S1–S3 examine measure, age, and segment dependencies; S4 reports exploratory prediction; S5–S7 assess processing stability, controlled signals, and model/partition sensitivity. S8 describes the retained external cohort and indexes the accompanying aggregate results archive.

**Unit of analysis.** CAUEEG entries are recordings, including repeated patients whose linkage is unavailable. All CAUEEG $p/q$ values, bootstrap intervals, and HC3 results below condition on independent recordings; they are retained for transparency and do not establish patient-level precision. Descriptive contrasts weight recordings, not unique patients. External ds004504 entries represent participants.

### S1 Complete default-measure comparison

All default measures are reported for both principal contrasts. Effect sizes use the sample-size-weighted pooled SD; tests use Welch's unequal-variance method. Raw $p$ and adjusted $q$ are shown separately. The correction family contains these 13 measures and the 29 distinct additional SE settings in each contrast; the duplicate default SE setting is counted once.

Table S1: Dementia versus normal: complete default-measure comparison on the same recordings ($n_{\text{disease}} = 306$, $n_{\text{normal}} = 457$). Welch $p$ and BH $q$ (42 measures) assume independent recordings and do not establish patient-level inference. AUC is direction-corrected.

| Measure | $d$ | $t$ | $p$ | $q$ | AUC |
|---|---|---|---|---|---|
| Sub-cycle PE | −0.690 | −9.37 | < 0.001 | < 0.001 | 0.699 |
| Cycle-spanning PE | −0.029 | −0.40 | 0.687 | 0.721 | 0.514 |
| Coarse-sampled PE | +0.705 | +9.61 | < 0.001 | < 0.001 | 0.701 |
| High-order PE | +0.009 | +0.13 | 0.900 | 0.921 | 0.504 |
| $SE_{\alpha}$ | +0.523 | +7.42 | < 0.001 | < 0.001 | 0.722 |
| $LZC_{\alpha}$ | +0.336 | +4.71 | < 0.001 | < 0.001 | 0.595 |
| LZC ratio | +0.473 | +6.45 | < 0.001 | < 0.001 | 0.624 |
| Relative alpha power | −0.568 | −8.02 | < 0.001 | < 0.001 | 0.660 |
| Relative theta power | +0.783 | +9.61 | < 0.001 | < 0.001 | 0.709 |
| Power ratio | −0.760 | −11.49 | < 0.001 | < 0.001 | 0.754 |
| PE ratio (o5d5) | +0.358 | +4.82 | < 0.001 | < 0.001 | 0.595 |
| PE ratio (o3d1) | −0.230 | −3.14 | 0.002 | 0.002 | 0.573 |
| SE ratio | +0.212 | +2.77 | 0.006 | 0.008 | 0.553 |

Table S2: MCI versus normal: complete default-measure comparison on the same recordings ($n_{\text{disease}} = 414$, $n_{\text{normal}} = 457$). Welch $p$ and BH $q$ (42 measures) assume independent recordings and do not establish patient-level inference. AUC is direction-corrected.

| Measure | $d$ | $t$ | $p$ | $q$ | AUC |
|---|---|---|---|---|---|
| Sub-cycle PE | −0.401 | −5.90 | < 0.001 | < 0.001 | 0.613 |
| Cycle-spanning PE | −0.034 | −0.50 | 0.614 | 0.661 | 0.512 |
| Coarse-sampled PE | +0.398 | +5.89 | < 0.001 | < 0.001 | 0.611 |
| High-order PE | −0.038 | −0.56 | 0.573 | 0.656 | 0.517 |
| $SE_{\alpha}$ | +0.258 | +3.82 | < 0.001 | < 0.001 | 0.598 |
| $LZC_{\alpha}$ | +0.315 | +4.66 | < 0.001 | < 0.001 | 0.594 |
| LZC ratio | +0.229 | +3.39 | < 0.001 | 0.001 | 0.560 |
| Relative alpha power | −0.293 | −4.34 | < 0.001 | < 0.001 | 0.575 |
| Relative theta power | +0.333 | +4.85 | < 0.001 | < 0.001 | 0.591 |
| Power ratio | −0.402 | −6.01 | < 0.001 | < 0.001 | 0.624 |
| PE ratio (o5d5) | +0.149 | +2.20 | 0.028 | 0.039 | 0.539 |
| PE ratio (o3d1) | −0.272 | −4.01 | < 0.001 | < 0.001 | 0.580 |
| SE ratio | +0.154 | +2.26 | 0.024 | 0.035 | 0.536 |

### S2 Age sensitivity

These analyses examine how the descriptive contrasts depend on the treatment of age differences between groups.

Table S3: Dementia-versus-normal age checks. Residualization uses all recordings; ages 70–80 includes 149 dementia and 129 normal recordings. Conditional $p/q$ assume independent recordings; BH covers 26 measure-by-approach tests. Patient-level precision is unresolved.

| | Raw | Age-residualized | | | Ages 70–80 | | |
|---|---|---|---|---|---|---|---|
| Measure | $d$ | $d$ | $p$ | $q$ | $d$ | $p$ | $q$ |
| Sub-cycle PE | −0.690 | −0.369 | < 0.001 | < 0.001 | −0.746 | < 0.001 | < 0.001 |
| Cycle-spanning PE | −0.029 | +0.030 | 0.677 | 0.677 | −0.292 | 0.017 | 0.027 |
| Coarse-sampled PE | +0.705 | +0.387 | < 0.001 | < 0.001 | +0.409 | < 0.001 | 0.001 |
| High-order PE | +0.009 | +0.053 | 0.460 | 0.520 | −0.242 | 0.047 | 0.065 |
| $SE_\alpha$ | +0.523 | +0.372 | < 0.001 | < 0.001 | +0.453 | < 0.001 | < 0.001 |
| $LZC_\alpha$ | +0.336 | +0.164 | 0.022 | 0.034 | −0.053 | 0.671 | 0.677 |
| LZC ratio | +0.473 | +0.367 | < 0.001 | < 0.001 | +0.232 | 0.056 | 0.072 |
| Relative alpha power | −0.568 | −0.430 | < 0.001 | < 0.001 | −0.420 | < 0.001 | 0.001 |
| Relative theta power | +0.783 | +0.475 | < 0.001 | < 0.001 | +0.577 | < 0.001 | < 0.001 |
| Power ratio | −0.760 | −0.486 | < 0.001 | < 0.001 | −0.640 | < 0.001 | < 0.001 |
| PE ratio (o5d5) | +0.358 | +0.280 | < 0.001 | < 0.001 | +0.125 | 0.298 | 0.369 |
| PE ratio (o3d1) | −0.230 | +0.058 | 0.429 | 0.507 | −0.267 | 0.028 | 0.040 |
| SE ratio | +0.212 | +0.236 | 0.002 | 0.004 | +0.077 | 0.523 | 0.566 |

The quadratic age term in normal recordings gave $p = 0.322$ and $q = 0.464$ for $SE_\alpha$, and $p = 0.605$ and $q = 0.715$ for the power ratio. Sub-cycle PE gave $p = 0.002$ and $q = 0.008$. The sub-cycle PE ratio changed from raw $d = -0.230$ to residualized $d = +0.058$ ($q = 0.507$). These checks illustrate sensitivity to age; neither adjustment establishes absence of confounding.

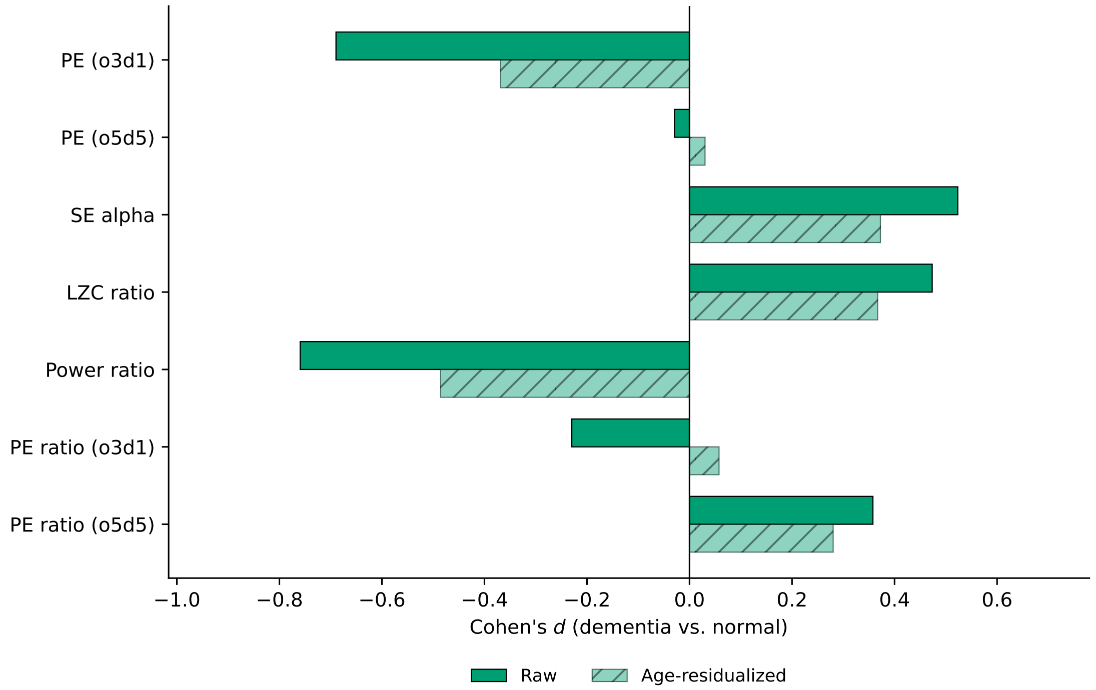


Figure S1: **Age adjustment changes effect magnitudes.** Paired bars show raw and linearly age-residualized pooled-SD Cohen's $d$. Residualization uses all recordings. Restriction to ages 70–80 is reported separately in Table S3; it is not individual matching.

### S3 SE parameter sensitivity and segment lengths

These checks separate finite estimator coverage from stability of the SE contrast under changes in parameters and segment length.

Whole-scalp effect ranges across tolerance were $d = -0.037$–$0.109$ for $m = 1$, 0.450–0.575 for $m = 2$, and 0.678–0.878 for $m = 3$. Minimum pairwise inter-tolerance correlations were 0.970, 0.739, and 0.666, respectively. Posterior-region ranges were $d = 0.093$–0.244, 0.552–0.664, and 0.729–0.918 for dimensions 1, 2, and 3. All 30 setting/region estimates were available for all 1,177 recordings; `baseline/group_statistics.csv` in `Supplementary_Results.zip` contains each effect, uncertainty interval, test, and group count.

Multiplying a nonconstant segment by a nonzero constant leaves SE unchanged when tolerance scales with that segment's SD. This invariance does not make the estimate insensitive to spectral content, dimension, tolerance, or segment length. The largest apparent AUC in the explored grid was 0.799; exploration on the full cohort makes this

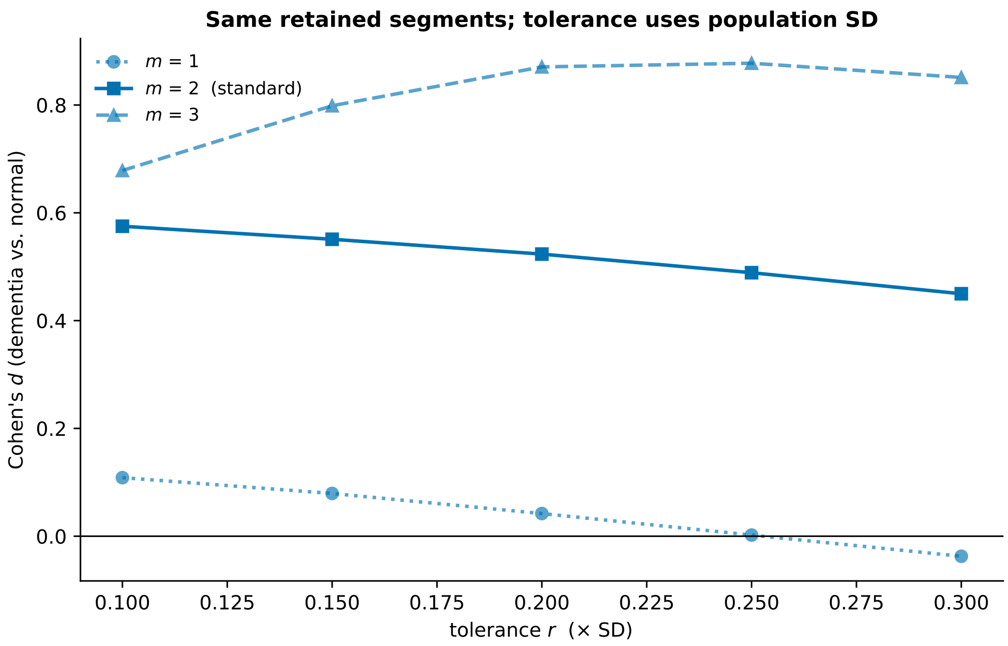


Figure S2: **SE sensitivity to dimension and tolerance on the same data.** Alpha-band dementia-versus-normal pooled-SD Cohen's $d$ is shown for $r = 0.10$–$0.30 \times$ SD at $m = 1, 2, 3$. All settings use identical retained segments and population SD, with complete finite estimator coverage. Dimension one compares scalar-template matches with two-sample extensions.

unsuitable as an unbiased estimate of a selected setting's future performance.

Table S4: Retained CAUEEG segment lengths in seconds. Segments are nested within recordings, with patient linkage unavailable; these are descriptive summaries, not independent observations for group testing.

| Group | Segments | Mean (s) | Median (s) | Range (s) |
|---|---|---|---|---|
| Dementia | 907 | 19.9 | 19.9 | 2.0–60.0 |
| MCI | 1417 | 17.0 | 12.4 | 2.0–60.0 |
| Normal | 1222 | 22.0 | 22.5 | 2.0–60.0 |

Entropy was estimated within individual segments, not on the total retained recording. Short segments can produce sparse matches or ordinal-state counts. Complete finite coverage resolves the earlier missing-estimate and partial-averaging problems but does not establish unbiased estimation. The descriptive segment distributions above do not establish that finite-sample effects are non-differential between groups. The completed duration, filtering, pooling, and synthetic checks are reported in Sections S5–S6; they show that complete coverage does not imply stable absolute estimates or group-effect magnitude.

### S4 Exploratory combined classifier

These analyses ask whether combining signal summaries improves discrimination under the stated partitions, with the exploratory and patient-grouping limitations detailed below.

For dementia versus normal, repeated stratified recording-level 10-fold cross-validation gave mean AUC 0.785 for power ratio plus $SE_\alpha$, with a split-percentile range of 0.781–0.789. Power alone gave 0.753 (0.751–0.756), and SE alone gave 0.722 (0.720–0.726). Adding the LZC ratio gave 0.795. The apparent two-feature AUC was 0.788.

These ranges are the 2.5th and 97.5th percentiles of ten repeat-mean AUCs from repeated partitions of the same recordings. They describe sensitivity to splitting and are not patient-sampling confidence intervals. Standardization was fit only in each training fold. Full-cohort feature selection preceded model cross-validation, and repeated patients may cross folds. Thus the analysis does not independently validate either the chosen feature set or performance on new patients.

For MCI versus normal, the two-feature model gave AUC 0.639, compared with 0.624 for power and 0.598 for SE. Adding age gave AUC 0.766 (split-percentile range 0.764–0.770), describing an exploratory association with age under recording-level splitting.

Interaction coefficients were tested in separate, unpenalized logistic models, rather than attaching ordinary likelihood standard errors to penalized coefficients. The power-by-SE term gave dementia $z = +0.37$, nominal $p = 0.711$, and MCI $z = -0.47$, nominal $p = 0.640$. Models with the interaction had mean cross-validated AUCs 0.785 and 0.639, respectively. These coefficient tests also assume independent recordings. Neither the interaction nor its recording-level cross-validation establishes patient-level or portable predictive value.

Table S5: Exploratory provider no-overlap train/test AUC. Test counts are disease/normal recordings. Feature development already used pooled data; within-partition repeated visits are unresolved.

| Contrast | Train $n$ | Test $n$ | Power | SE | Power + SE |
|---|---|---|---|---|---|
| Dementia | 610 | 22/35 | 0.690 | 0.742 | 0.740 |
| MCI | 696 | 33/35 | 0.472 | 0.660 | 0.524 |

The provider's `dementia-no-overlap.json` removes validation/test recordings from patients represented in training (`https://github.com/ipis-mjkim/caueeg-dataset`). Training was unchanged; only the official test partition was evaluated, without pooling validation and test. Standardization was fitted on training only; logistic regression used fixed default L2 regularization ($C = 1$). Features had already been developed using pooled records, and within-partition repeated patients remain unidentified. This is an exploratory transfer check, not untouched validation. The combination showed no established advantage over SE alone in dementia; MCI results were less consistent. AUCs here retain the trained direction.

### S5 Duration, filtering, and aggregation sensitivity

These comparisons test whether the opposing PE directions and alternative contrasts persist when signal preparation changes. Matched eligible subsets distinguish processing from sample-composition changes.

All 1,177 CAUEEG recordings and 78 external participants were processed under an exploratory protocol fixed before complete sensitivity tables were inspected. The original values reproduced to relative tolerance $10^{-12}$ and absolute tolerance $10^{-13}$; 168,113 windows passed sample-boundary checks, with zero extraction errors. The protocol is not a formal preregistration. Each variant retained original channels and accepted clean intervals. Nonoverlapping windows never crossed interval boundaries; unused remainders were discarded. Windows of 2, 4, 8, or 16 s were selected after parent-segment filtering. Two 2 s specifications instead filtered each window separately. The interior pair selected windows at least 165 samples (0.825 s, half the 331-tap FIR span) inside each parent boundary. Hilbert transforms used the analysis window in both filtering conditions. Interior selection reduces direct parent-boundary support but does not remove window-boundary effects from every estimator.

Variants required at least 10 s of whole windows among originally eligible records; the fixed-budget variant used exactly the first ten 2 s windows (20 s). No excluded recording was added. Group summaries required at least 20 eligible records per group. Every processing comparison to baseline used the same eligible records. `robustness/effects_and_paired_changes.csv` in `Supplementary_Results.zip` retains paired recording-bootstrap results (2,000 draws, seed 20260925) and 17-measure BH families per variant/contrast; these are conditional on recording independence and do not correct across exploratory specifications.

Table S6: CAUEEG dementia versus normal: descriptive $d$ under nine specifications. Sub, Cycle, Coarse, and High denote the four PE representations; Power is the alpha/theta power ratio. $n$ is disease/reference (recordings in CAUEEG; participants externally). Dashes indicate fewer than 20 eligible entries in a group. Original effects on the same eligible subsets and all pooled/quality-exclusion results are in `robustness/effects_and_paired_changes.csv` in `Supplementary_Results.zip`.

| Specification | $n$ | Sub | Cycle | Coarse | High | SE | LZC ratio | Power |
|---|---|---|---|---|---|---|---|---|
| Original | 306/457 | −0.690 | −0.029 | +0.705 | +0.009 | +0.523 | +0.473 | −0.760 |
| 2 s, window | 306/457 | −0.606 | +0.020 | +0.703 | +0.084 | +0.205 | +0.291 | −0.742 |
| 2 s, parent | 306/457 | −0.692 | +0.016 | +0.712 | +0.078 | +0.197 | +0.260 | −0.742 |
| 4 s, parent | 306/456 | −0.671 | +0.020 | +0.703 | +0.065 | +0.366 | +0.353 | −0.752 |
| 8 s, parent | 301/446 | −0.709 | +0.009 | +0.701 | +0.051 | +0.489 | +0.412 | −0.753 |
| 16 s, parent | 291/435 | −0.729 | −0.014 | +0.685 | +0.029 | +0.435 | +0.451 | −0.766 |
| 2 s interior, window | 305/456 | −0.576 | +0.030 | +0.693 | +0.091 | +0.138 | +0.282 | −0.759 |
| 2 s interior, parent | 305/456 | −0.685 | +0.023 | +0.709 | +0.082 | +0.120 | +0.250 | −0.759 |
| 2 s, exactly 20 s | 305/456 | −0.716 | −0.026 | +0.625 | +0.031 | +0.220 | +0.197 | −0.700 |

Table S7: External AD versus control: descriptive $d$ under nine specifications. Sub, Cycle, Coarse, and High denote the four PE representations; Power is the alpha/theta power ratio. $n$ is disease/reference (recordings in CAUEEG; participants externally). Dashes indicate fewer than 20 eligible entries in a group. Original effects on the same eligible subsets and all pooled/quality-exclusion results are in `robustness/effects_and_paired_changes.csv` in `Supplementary_Results.zip`.

| Specification | $n$ | Sub | Cycle | Coarse | High | SE | LZC ratio | Power |
|---|---|---|---|---|---|---|---|---|
| Original | 31/27 | +0.245 | +0.755 | +1.159 | +0.873 | +0.011 | +0.495 | −0.940 |
| 2 s, window | 31/27 | +0.275 | +0.677 | +1.125 | +0.759 | −0.295 | +0.516 | −0.915 |
| 2 s, parent | 31/27 | +0.244 | +0.661 | +1.139 | +0.736 | −0.174 | +0.487 | −0.915 |
| 4 s, parent | 27/21 | +0.337 | +0.705 | +1.035 | +0.747 | −0.291 | +0.269 | −0.636 |
| 8 s, parent | – | – | – | – | – | – | – | – |
| 16 s, parent | – | – | – | – | – | – | – | – |
| 2 s interior, window | – | – | – | – | – | – | – | – |
| 2 s interior, parent | – | – | – | – | – | – | – | – |
| 2 s, exactly 20 s | 31/27 | +0.310 | +0.668 | +0.957 | +0.725 | −0.172 | +0.506 | −0.908 |

Table S8: CAUEEG: eligible counts and median analyzed seconds; entries follow Dementia, MCI, Normal. Original eligibility is retained, and incomplete whole windows are discarded.

| Specification | Eligible | Median seconds |
|---|---|---|
| Original | 306/414/457 | 60/60/60 |
| 2 s, window filter | 306/414/457 | 58/58/58 |
| 2 s, parent filter | 306/414/457 | 58/58/58 |
| 4 s, parent filter | 306/414/456 | 56/56/56 |
| 8 s, parent filter | 301/401/446 | 48/48/48 |
| 16 s, parent filter | 291/375/435 | 32/32/32 |
| 2 s interior, window | 305/412/456 | 54/54/54 |
| 2 s interior, parent | 305/412/456 | 54/54/54 |
| 2 s, exactly 20 s | 305/412/456 | 20/20/20 |

Table S9: EXTERNAL: eligible counts and median analyzed seconds; entries follow AD, Control, FTD. Original eligibility is retained, and incomplete whole windows are discarded.

| Specification | Eligible | Median seconds |
|---|---|---|
| Original | 31/27/20 | 60/60/60 |
| 2 s, window filter | 31/27/20 | 54/54/53 |
| 2 s, parent filter | 31/27/20 | 54/54/53 |
| 4 s, parent filter | 27/21/16 | 20/20/20 |
| 8 s, parent filter | 4/1/2 | 16/16/20 |
| 16 s, parent filter | 2/0/1 | 16/–/16 |
| 2 s interior, window | 21/14/11 | 16/14/16 |
| 2 s interior, parent | 21/14/11 | 16/14/16 |
| 2 s, exactly 20 s | 31/27/20 | 20/20/20 |

The dementia sub-cycle/coarse directions persisted across all nine specifications. Window filtering modestly attenuated sub-cycle d; excluding records with constant windows restored much of that difference. SE fell from $d = +0.523$ to $+0.197$ on parent-filtered 2 s windows using the same 306/457 dementia/normal records, and to $+0.120$ for interior windows. Thus SE magnitude was not invariant to segment definition. External SE changed from $d = +0.011$ originally to $-0.174$ with parent-filtered 2 s windows; a near-zero original estimate is not evidence of equivalence or an established cohort difference.

Constant-window flags affected three CAUEEG normal recordings under window filtering and one under parent filtering. Dropping every flagged recording preserved the main PE reversal. The largest d change was sub-cycle PE in the window-filtered interior contrast, from $-0.576$ to $-0.698$; its SE effect fell from $+0.138$ to $+0.092$. All included-record summaries remained finite under the declared constant-signal convention.

Ordinal counts were pooled within each channel and recording without vectors crossing window boundaries, then normalized by $\log(m!)$. Pooling and mean segment entropy answer different questions: pooled entropy can include differences in pattern distributions between segments. In CAUEEG, mean high-order entropy was 0.579 originally and 0.534 with parent-filtered 2 s windows, whereas pooled-count values were 0.585 and 0.585. The high-order dementia contrast remained small under both rules; the pooled fixed-20 s estimate was $d = -0.008$. Neither aggregation is established as universally unbiased.

### S6 Controlled signals and spectral probes

Controlled generators and spectral probes examine which measurement behaviors can occur without a disease process, establishing possible signal accounts of the observations.

Synthetic signals were sampled at 200 Hz. Frequency sweeps covered 8–12 Hz in 0.1 Hz steps, eight phase offsets, and alpha-noise SD 0, 0.01, or 0.1. Length curves used 50 seeded realizations each of a 10 Hz sinusoid, a sinusoid plus filtered noise, and alpha-filtered Gaussian noise, with nested 2/4/8/16/32/60 s prefixes. Alpha-filtered noise showed approximately 88.5% phase-pattern agreement without a disease process. Its high-order entropy increased from approximately 0.565 at 2 s to 0.629 at 60 s; averaging 2 s entropies stayed near 0.568, while pooling their counts approached 0.629. These are generator-specific demonstrations of estimation and aggregation sensitivity, not a universal bias correction.

For noisy sinusoids, sub-cycle PE rose with frequency while coarse PE had a trough near 10 Hz. Slowing below 10 Hz can therefore generate opposing changes in these two settings. The analytical uniform-phase sector calculation gave normalized entropy 0.607; it averages over starting phase, rather than assuming a single commensurately sampled periodic orbit covers that continuum. These checks support a signal-geometric account without proving a unique clinical mechanism.

The spectral probe selected 20 CAUEEG recordings per diagnostic group and all 58 external AD/control participants, with ten independent Fourier-phase randomizations

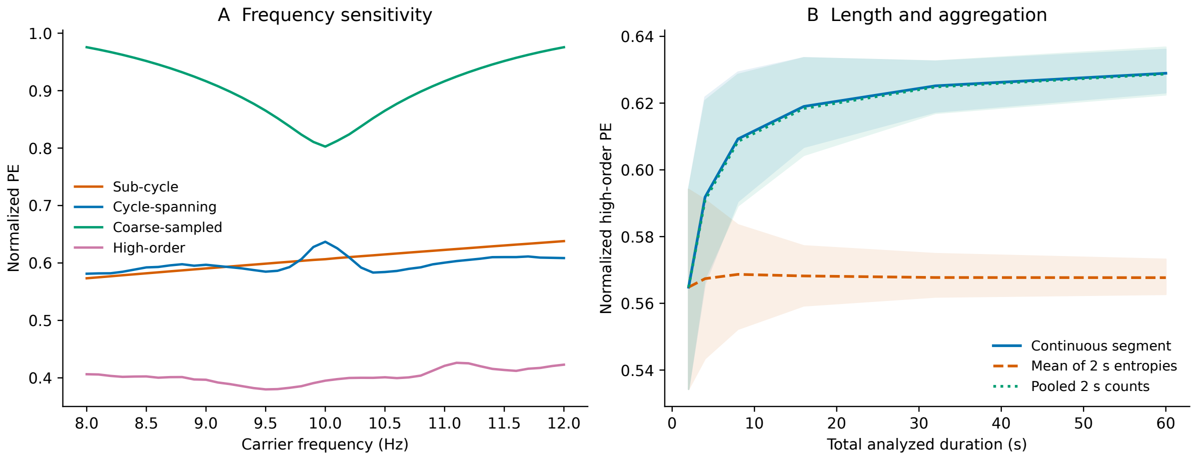


Figure S3: **Controlled signals reproduce frequency and length sensitivity.** Left: mean PE across eight phase offsets for sinusoids with alpha-filtered noise (SD 0.1), using the four tested representations. Coarse PE has a trough near 10 Hz while sub-cycle PE rises. Right: mean high-order PE across 50 alpha-filtered Gaussian-noise realizations, using nested durations. Shading spans the 2.5th–97.5th percentiles of simulated values, not a confidence interval for clinical effects. Pooling counts across 2 s windows approaches continuous-segment entropy while averaging window entropies remains lower. No vectors cross window boundaries. These are known-generator demonstrations, not disease models.

per recording. Each channel/clean parent segment retained its Fourier magnitudes and mean. Circular-boundary assumptions and changed amplitude distributions limit interpretation; no formal surrogate test of nonlinearity was performed. CAUEEG phase-pattern agreement was 90.24% observed and 90.13% after randomization. On the same 20 dementia/20 normal probe records, sub-cycle d changed from −0.343 to −0.234 and coarse d from +0.485 to +0.575 when each record was represented by its ten-surrogate mean. Averaging surrogates changes the distribution and reduces Monte Carlo variability, so these d values are descriptive. High phase reconstruction alone does not establish disease-specific nonlinear complexity.

### S7 Flexible age models and partition sensitivity

These checks assess whether the descriptive representation contrast depends on the chosen age model or provider partition.

Models used all CAUEEG recordings with linear age, quadratic age, a centered natural cubic spline (four degrees of freedom), or group-specific splines. Fitted diagnostic contrasts were averaged over observed ages 70–80 (508 recordings) and expressed in full-cohort outcome SD, not Cohen's d. This standardization does not establish individual matching or causal adjustment. The HC3 and 13-measure BH outputs in `age/age_shape_contrasts.csv` do not account for repeated patients or specification search.

Table S10: Joint age-model contrasts standardized over observed ages 70–80, in full-cohort outcome SD. All fits use 1,177 CAUEEG recordings; these are model-dependent associations, not patient-adjusted or causal effects.

| Model | Dem. sub-cycle | Dem. coarse | MCI sub-cycle | MCI coarse |
|---|---|---|---|---|
| Linear | −0.454 | +0.482 | −0.230 | +0.221 |
| Quadratic | −0.444 | +0.480 | −0.234 | +0.222 |
| Spline | −0.434 | +0.481 | −0.214 | +0.237 |
| Group-specific spline | −0.611 | +0.320 | −0.280 | +0.046 |

Dementia sub-cycle/coarse directions persisted; MCI coarse contrasts were small under group-specific curves. Removing 65 provider-identified train-overlap recordings left 1,112 records, with dementia contrasts changing modestly (sub-cycle −0.690 to −0.681; coarse +0.705 to +0.719). This union is not patient-deduplicated. Official test contrasts were −1.081 and +0.647 for dementia (22/35 records), and −0.128 for MCI coarse PE (33/35). Validation dementia sub-cycle d was −0.005 (19/34). Small partitions and model sensitivity limit replication claims. Patient linkage is needed for clustered uncertainty and prespecified visit selection.

### S8 Retained external cohort and aggregate results

The external summaries below describe the 78 participants retained after processing. The reported AD–control contrast uses 31 AD and 27 control participants; the remaining 20 had FTD. Gender is reported using the source metadata labels F and M, without inferring whether the field represents sex or gender identity. It was not included as a model covariate. Comparable severity and sex/gender summaries were unavailable for CAUEEG.

Table S11: Retained ds004504 cohort. Age and MMSE are mean ± sample SD. Gender entries are source-labeled F/M counts; none were missing. Channels are median (range) after exclusion.

| Group | $n$ | Age (years) | MMSE | F/M | Channels |
|---|---|---|---|---|---|
| AD | 31 | $67.1 \pm 8.1$ | $17.5 \pm 4.8$ | 21/10 | 19 (11–19) |
| Control | 27 | $68.1 \pm 5.5$ | $30.0 \pm 0.0$ | 10/17 | 18 (11–19) |
| FTD | 20 | $64.8 \pm 7.9$ | $22.0 \pm 2.7$ | 8/12 | 19 (11–19) |

Table S12: External AD versus control: 31 AD and 27 control participants. Intervals are pointwise 95% participant-bootstrap intervals; $p$ is Welch and $q$ is BH across 13 defaults. Single-measure AUC is direction-corrected after observing direction.

| Measure | $d$ | 95% interval | $p$ | $q$ | AUC |
|---|---|---|---|---|---|
| Sub-cycle PE | +0.245 | [−0.321, +0.740] | 0.359 | 0.424 | 0.514 |
| Cycle-spanning PE | +0.755 | [+0.270, +1.277] | 0.008 | 0.018 | 0.691 |
| Coarse-sampled PE | +1.159 | [+0.662, +1.808] | $<$ 0.001 | 0.001 | 0.815 |
| High-order PE | +0.873 | [+0.375, +1.400] | 0.003 | 0.008 | 0.722 |
| $\mathrm{SE}_\alpha$ | +0.011 | [−0.432, +0.659] | 0.966 | 0.966 | 0.600 |
| LZC ratio | +0.495 | [+0.005, +1.059] | 0.071 | 0.115 | 0.631 |
| Power ratio | −0.940 | [−1.499, −0.495] | 0.001 | 0.006 | 0.802 |
| Relative alpha power | −0.812 | [−1.328, −0.375] | 0.006 | 0.014 | 0.718 |
| Relative theta power | +0.871 | [+0.404, +1.423] | 0.001 | 0.006 | 0.736 |
| $\mathrm{LZC}_\alpha$ | +0.254 | [−0.192, +0.864] | 0.337 | 0.424 | 0.619 |
| PE ratio (o5d5) | +0.692 | [+0.187, +1.263] | 0.014 | 0.025 | 0.680 |
| PE ratio (o3d1) | +0.419 | [−0.094, +0.961] | 0.125 | 0.180 | 0.590 |
| SE ratio | +0.229 | [−0.260, +0.769] | 0.393 | 0.426 | 0.564 |

The accompanying `Supplementary_Results.zip` provides machine-readable aggregate results. Its `README.md` defines the fields, analysis populations, and inference limitations; `manifest.json` identifies the frozen code version, source runs, and file hashes. It contains no EEG, recording identifiers, participant-level clinical rows, or individual predictions. The file locations below are relative to that archive.

- `baseline/group_statistics.csv`: the 13 defaults and 29 additional SE settings for each CAUEEG contrast; `baseline/external_statistics.csv`: all external default contrasts and participant-bootstrap intervals.
- `baseline/age_statistics.csv` and `baseline/age_nonlinearity.csv`: age-model results. Phase, frequency-correlation, and centroid-model summaries are in the same folder; see the archive's file map.
- `baseline/cross_validation.csv` and `baseline/interaction.csv`: exploratory

recording-level prediction and interaction summaries.

- `robustness/effects_and_paired_changes.csv`: all reported sensitivity measures, including all four PE settings, pooled-count variants, matched-eligibility original effects, and constant-window exclusions. `robustness/eligibility.csv` reports counts and duration; the other files in this folder report phase, surrogate, representation-gap, and age-model summaries.
- `age/age_shape_contrasts.csv` and `partitions/`: flexible-age and provider-partition aggregate results.
- `simulations/`: the frequency-sweep and duration-curve outputs underlying Figure S3, with simulation settings and the analytical phase check.
- `cohorts/external_cohort_summary.csv`: retained external participant characteristics corresponding to Table S11.

All CAUEEG inferential outputs remain conditional on recording independence. Resampling preserves pairing between measures or specifications on one recording, but cannot group repeated visits from the same patient. Cross-validation percentile ranges describe repeated splits, and synthetic percentile bands describe simulated realizations. Neither is a patient-sampling confidence interval.
